# Reducing placebo response in clinical trials of agitation in Alzheimer’s disease

**DOI:** 10.64898/2026.06.03.26354808

**Authors:** Karin C. Knudson, Kevin M. Anderson, Michael Ballard, Robert A. Lenz, Tien Dam, Doron Sagman, Nicholas J. Brandon, Tathagata Banerjee, Andrew E. Jaffe

**Affiliations:** Neumora Therapeutics, Inc. Watertown MA, 02472

## Abstract

High placebo response is an obstacle in developing drugs to treat agitation in Alzheimer’s disease (AAD), a prevalent and burdensome symptom. However, it has proved challenging to develop actionable models of placebo response that 1) can be applied prospectively, requiring only information available at screening or baseline, 2) yield strategies for reducing placebo response without equally depressing drug response, and 3) show generalizability across trials. Here, we first investigated placebo response in AAD at the trial level using meta-regression applied to 23 clinical trials. Meta-regression identified several factors associated with increased placebo response, but most of these factors were non-specific such that they predicted improvements in drug response as well. We therefore turned to individual level clinical trial datasets and applied causal modeling to predict which participants would have high placebo response relative to predicted drug response. We successfully built and validated the causal model across two independent clinical trials of risperidone and haloperidol at the level of individual patients (ability to predict subsequent improvement on drug or placebo). Crucially, we also found efficacy improvements in the overall trial through *in silico* exclusion/screen failing of high placebo-predicted subjects. We further characterized features most associated with placebo response to improve explainability and, lastly, validated the effect of these features at the trial level in clinical trials of galantamine, an acetylcholinesterase inhibitor (hence in a different class of drugs than those in the other two trials used). Taken together, we have developed and applied a causal modeling framework for reducing placebo response and increasing trial-level efficacy in neuropsychiatry clinical trials using historical trial datasets.

## Introduction

Alzheimer’s disease (AD) is a common and devastating neurodegenerative disorder that primarily affects cognition and memory with accompanying neuropsychiatric symptoms such as psychosis, depression, irritability, and/or aggression^1^. These accompanying behavioral and psychiatric symptoms of dementia (BPSD) are common in AD populations, with a prevalence measured above 90% ^2,3^ .Specific prevalence estimates of agitation range from 24% to 88%^4–6^. Given the high global occurrence of AD dementia (∼32 million) and prodromal or preclinical AD (∼384 million)^7^, combined with increasing incidence—doubling every 20 years^8^ —comorbid neuropsychiatric symptoms in AD represent a huge global burden.

BPSD, which include agitation, are not only highly prevalent, but are also associated with undesirable outcomes for patients, caregivers, and the broader community. For patients, BPSD are associated with higher rates of mortality^9^, faster progression to severe AD and death^10^, and decreased quality of life^11,12^. For caregivers, the severity of a patient’s BPSD—and not cognitive deficits—is associated with higher caregiver burden^13,14^ and decreased caregiver quality of life^15^. Agitation in AD is solely associated with caregiver burden and caregiver depression, after controlling for other factors^2^. Agitation in AD is further associated with an elevated risk of patient institutionalization, thereby incurring billions in estimated incremental cost.^16^

Treatments for agitation in AD include nonpharmacologic interventions as a first line approach, followed by use of antipsychotics, sedative/hypnotics, anxiolytics, acetylcholinesterase inhibitors, memantine, and antidepressants^17^. These treatment options are often unsatisfactory in the balance of efficacy and risk; atypical antipsychotics are pervasive but have been associated with adverse events and an increased risk of death^18,19^. For example, a recent large cohort study of adults with dementia (N=173,910) found antipsychotic use is associated with a broad set of adverse outcomes: stroke, venous thromboembolism, myocardial infarction, heart failure, fracture, pneumonia, and acute kidney injury^20^. Brexpiprazole is approved for the treatment of agitation associated with dementia due to Alzheimer’s disease, but has a boxed warning for increased mortality in elderly patients with dementia-related psychosis. While the very recent approval of dextromethorphan-bupropion (April 2026) has expanded the available options, significant unmet need remains.

Given the high prevalence and negative impact of BPSD and agitation in AD, and downsides associated with current treatments, there is an urgent need to develop better treatments. However, one of the largest barriers to developing new treatments has been robust placebo responses within clinical trials for new therapies^21^. A meta-analysis of 13 RCTs for treatment of BPSD between 2009 and 2015 found statistically significant improvement within the placebo groups of on average –2.68 points on the Neuropsychiatric Inventory (NPI), approaching clinically meaningful improvement^22^.

Moreover, the variation in placebo response between studies is great, so individual studies can have much higher placebo responses than similar trials. Unexpectedly high placebo response was specifically cited as the reason for lack of significance in a 24-week trial of memantine for AD^23^, and in trials with BPSD focused endpoints, improvement on placebo has sometimes numerically exceeded improvement on drug^24^. Furthermore, the challenge of addressing placebo response is growing; meta-analysis has shown that the placebo effect is increasing over time in BPSD, as it has in other psychiatric areas^22^. Understanding key drivers of placebo response in this patient population—and subsequently leveraging these factors to guide clinical development to reduce placebo response in future trials—could transform the landscape of treating agitation in AD and improve the lives of patients and their caregivers.

Here we first characterize the extent of placebo response in clinical trials of agitation in AD by applying meta-regression using published summary statistics. We next use subject-level data from clinical trials of risperidone, a second generation antipsychotic, to build and then independently validate a placebo response model of agitation using causal inference methodologies. We demonstrate that our model can predict placebo response at the level of individual patient (e.g. accurate predictions of outcome improvements when randomized to placebo) and overall trial (e.g. increased/enriched treatment effect when removing predicted placebo-responsive subjects). We lastly characterize clinical factors associated with placebo response and demonstrate generalizability across mechanisms of the placebo response model to enhance treatment effects within studies of galantamine, an acetylcholinesterase inhibitor. These results together suggest that a small fraction of patients scoring high on symptom scales at baseline disproportionately drive placebo response in studies of agitation in AD, and that excluding such patients could increase placebo-adjusted efficacy of treatments.

## Methods

### Meta-regression data collection, harmonization and analysis

#### Search strategy and study selection criteria

To conduct the search, one author (KMA) searched PubMed, clinicaltrials.gov, and Google Scholar for clinical trials of AAD published until August 28, 2023. The following eligibility criteria were applied to filter studies to those that a) had an active placebo arm, b) had assessment of treatment effect on agitation or agitation/aggression as an objective, and c) did not target psychosis as a primary endpoint, and d) included the Cohen Mansfield Agitation Inventory (CMAI)^25^ or Neuropsychiatric Inventory agitation/aggression domain score (NPI-AA) as clinical endpoints. No automation tools were used to identify studies or extract data.

#### Data collection and features

We manually extracted baseline characteristics, study features, and treatment effects within the active and placebo arms from the main tables in each publication. Study-level features included: Year, Number of Treatment Weeks, Number of Sites, Visit Interval, Length of Study Lead-in, Number of Trial Arms, Percent Allocated to Placebo, and Enrollment Age Minimum. Arm-level (participant summary) features included: Mean Age, Percent Female, Percent in Care Facility, CMAI Baseline, NPI-AA Baseline, MMSE Baseline, NPI Caregiver Burden Baseline, and Mean Months Since AD Diagnosis. Primary outcomes included change from baseline on the CMAI and NPI-AA endpoints.

#### Effect measures and harmonization

Outcome measures were standardized change score effect sizes (i.e. Cohen’s d_z_) on the CMAI and NPI-AA endpoints. For studies that only reported mean values at study endpoint, we manually calculated the change from baseline score. Studies that reported both CMAI and NPI-AA showed high convergence of standardized change scores of CMAI and NPI-AA (r=0.74, Figure S1).

Raw variance estimates were reported either as the standard deviation (SD), confidence intervals, or standard error. Variance estimates were harmonized to standard error of change values for use in meta-regression. When only the variance of the endpoints was available, we imputed the variance of the changes scores using the following formula, which assumes 0.6 for the correlation coefficient between participants’ baseline and final measurements based on previous studies^26–28^:

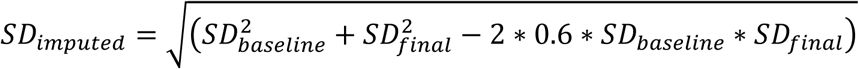

^29,30^. When multiple variations of the clinical scales were present, values were rescaled linearly to match the more common range within the studies (e.g. some studies used the 36-item version of the for CMAI with each item rated 0-6 leading to a possible CMAI range of 0-216; total scores were rescaled to the more common 29-203 range). When a trial had multiple drug arms, the arm with the greatest effect size was selected for comparison.

We expect heterogeneity in effect size due to the different treatments, study duration, and other study features. While publication bias is difficult to assess in the presence of such heterogeneity, Egger’s regression test did not detect significant funnel plot asymmetry (p=0.22, Figure S2).

#### Analysis

Meta-analysis of placebo arm changes was conducted using the meta package (v. 8.2-1) in R with a random effects model and Hartung-Knapp (HK) adjustment. Meta-regression to assess study- and arm-level covariates was conducted with the PyMare package (v.0.0.10) in Python. For each study and arm-level covariate, we fit an independent meta-regression model across all available studies first for the drug arms and then for the placebo. The metaregression models were fit using the Restricted Maximum Likelihood (REML) approach. A model was only fit if there were at least five arms within the relevant group of drug or placebo arms for which the feature was available. Adjusted p-values are reported, correcting for multiple hypotheses using the Benjamini-Hochberg (BH) correction.

### Subject-level datasets

We analyzed historical clinical trial datasets of small molecules used to treat symptoms of Alzheimer’s disease from the Yale University Open Data Access Project under project/proposal #2023-5295. These trials were conducted by Janssen Research & Development in the 1990s. We specifically used the “RIS-AUS-5" trial (NCT00249158)^31^ as the dataset used to train the placebo response model. This trial was selected as the training dataset based on the presence of the Cohen Manfield Agitation Inventory (CMAI)^25^ at baseline and follow-up, and presence of a single drug arm (risperidone) and a placebo arm. We used the “RIS-INT-24" trial (NCT00249145)^32^ of risperidone, haloperidol and placebo as the held-out testing dataset which had a similar patient population and measured similar clinical scales (Figure S3, S4).

We also used two trials of galantamine on cognitive and behavioral endpoints— GAL-USA-10”^33^ and “GAL-INT-10"^34^—to explore the generalizability of the placebo response models to drug classes beyond antipsychotics in AD patient populations. These two trials were selected as comparison datasets because alongside their cognitive primary efficacy endpoints, they also measured BPSD (in both cases with the Neuropsychiatric Inventory, NPI). Moreover, their large study sizes meant that although the trials did not enroll patients based on presence or severity of agitation, many patients had clinically significant agitation and aggression at baseline (via NPI-AA ≥ 4), including (125 in GAL-USA-10 (13%) and 95 in GAL-INT-10 (10%)). In our analysis of the galantamine datasets, we pooled the drug arms from the two studies, which meant combining drug arms with differences in dosing.

### Placebo Response Model Training

From the N=326 subjects in RIS-AUS-5 that had CMAI measurements at both baseline and the 12 week follow-up visit (or date of discontinuation, as was used in the original study if 12 week data was missing), we used the subset of N=305 subjects with complete data on the following clinical and demographic information at baseline (or screening if baseline values were not available, as for the MMSE and demographic variables): the Mini Mental State Examination (MMSE)^35^, Behavioral Pathology in Alzheimer’s Disease Rating Scale (BEHAVE-AD)^36^ total and sub-scores (agitation/aggressiveness, affective disturbances, diurnal disturbances, paranoid and delusional ideation, hallucinations, activity disturbances, and anxiety) , CMAI total and CMAI subscores (verbal aggressive , physical aggressive, verbal nonaggressive, physical nonaggressive), age, sex, and clinical symptomatology related to depression, delusions, and vascular type. In the only exception to requiring complete baseline training data for inclusion, we included the 40 participants in the training dataset who were missing the two individual BEHAVE items corresponding to delusions about abandonment and one’s house not being one’s home, filling the missing values with 0, the modal values for those items (70% and 80% percent of non-missing baseline scores were 0 for each of these two items respectively). Features were standard-scaled before model-fitting.

We used a Linear XLearner method^37^ as the causal model that was trained on the RIS-AUS-5 dataset, with 5-fold cross-validated linear elastic nets as the base potential outcome models, and an l2-regularized logistic regression model as the propensity model. Like other causal models, the XLearner, when applied to a baseline profile from a patient, produces a patient-level conditional average treatment effect (CATE), which is the predicted expected difference in the potential outcome (here, change in CMAI) if the patient were given the active compound and the potential outcome if the participant were given the placebo. The choice of linear, regularized, X-Learner model as the form of our “CMAI Placebo Response Model” was suitable due to its high interpretability and its ability to estimate potential outcomes alongside treatment effect, From it we obtain highly interpretable (linear) placebo and drug response models as components of the overall model. Moreover, the model form encourages sparsity so that coefficients for clinical characteristics that are irrelevant to the potential outcomes are pushed to zero.

### Placebo Response Model Validation

The placebo response model trained on the RIS-AUS-5 dataset (risperidone) was then applied to the baseline features from the independent RIS-INT-24 dataset (risperidone and haloperidol) to produce the XLearner’s predicted CATE values for each new/unseen participant. The RIS-INT-24 baseline covariates were scaled using the standard scaler that was fit on the training RIS-AUS-5 data (i.e. subtracting the training data mean from each of the covariate values and dividing by the training data standard deviation of each covariate). To assess the study-level generalizability of the placebo response model, various study-level metrics were considered with and without dropping the participants with the highest predicted CATE values (i.e. those with the strongest placebo response relative to their drug response). For instance, we exclude those predicted to be in the top 5% of CATE, and compute for those in the remaining 95% the mean improvement on placebo, mean improvement on drug, the treatment Cohen’s d effect size, and the significance of the treatment effect on change in CMAI in an ANCOVA controlling for baseline severity. Where percentile cutoffs were used to assess the effect of omitting patients, the percentile cutoffs were calculated using the training data (see Table S2 for the CATE values that corresponded to each percentile). This style of study-level assessment allows for comparison of these study-level results as computed with all participants to the results when participants are excluded using the model estimates computed from their baseline characteristics. Therefore, it is directly related to the effect on the study of prospectively applying a model-based inclusion/exclusion criterion.

In addition to this study-level validation of the model, we also consider patient-level accuracy. X-Learner models include at their first level two separately fit models that predict drug and placebo potential outcomes, which feed into the conditional average treatment effects for controls/treated that are combined with a fitted propensity model to produce CATE estimates. To assess the patient-level accuracy of the underlying potential outcome models trained in the first stage of the XLearner, these two linear models were used to predict the outcome (change in CMAI) of the validation dataset drug and placebo arm participants respectively, so that the true and predicted outcomes could be compared within each arm.

### Efficacy Analysis Methods

Treatment effects were calculated using linear regression, with the response variable as the 12-week change in total CMAI (with last observation carried forward, LOCF), the treatment variable was binarized drug or placebo, and baseline total CMAI was included as a covariate. We also report the Cohen’s d as a standardized measure of treatment effect size.

## RESULTS

### Quantifying placebo response in clinical trials of agitation in Alzheimer’s Disease

We identified 23 clinical trials – consisting of 55 trial arms (23 placebo arms and 32 active arms) that met our criteria of having: a) an active placebo arm, b) enrolling patients with elevated behavioral agitation, c) not targeting psychosis as a primary endpoint, and d) including CMAI or NPI-AA as clinical endpoints (Table S2). We standardized effect sizes for the change-from-baseline efficacy measures in each arm in each trial (see Methods) – weighted meta-analysis across the 23 placebo arms indicated significant effects of placebo (d= -0.630, p < 0.0001).

Meta-regression with study-level factors in both drug and placebo arms identified that the number of sites, mean baseline severity, year of study, number of visits, and lower mean age were significantly (p-value < 0.05 after Benjamini-Hochberg correction) associated with larger improvements from baseline in agitation symptoms in the placebo arms across these trials. (Figure 1B). However, with the exception of age (drug arms adj. p-value=0.14) all of these predictors significantly associated with increasing response to active treatment in these same trials, suggesting that modifying trial criteria based on many of the factors identified in analyses of historical placebo arms likely would reduce changes in outcomes in the active arms as well. Together these analyses quantify the magnitude of placebo response across recent trials of agitation in AD, and also show the challenges of using study-level inference to identify clinical factors that distinguish placebo response from drug response, especially when such factors can affect both drug and placebo response simultaneously.

**Figure 1:**
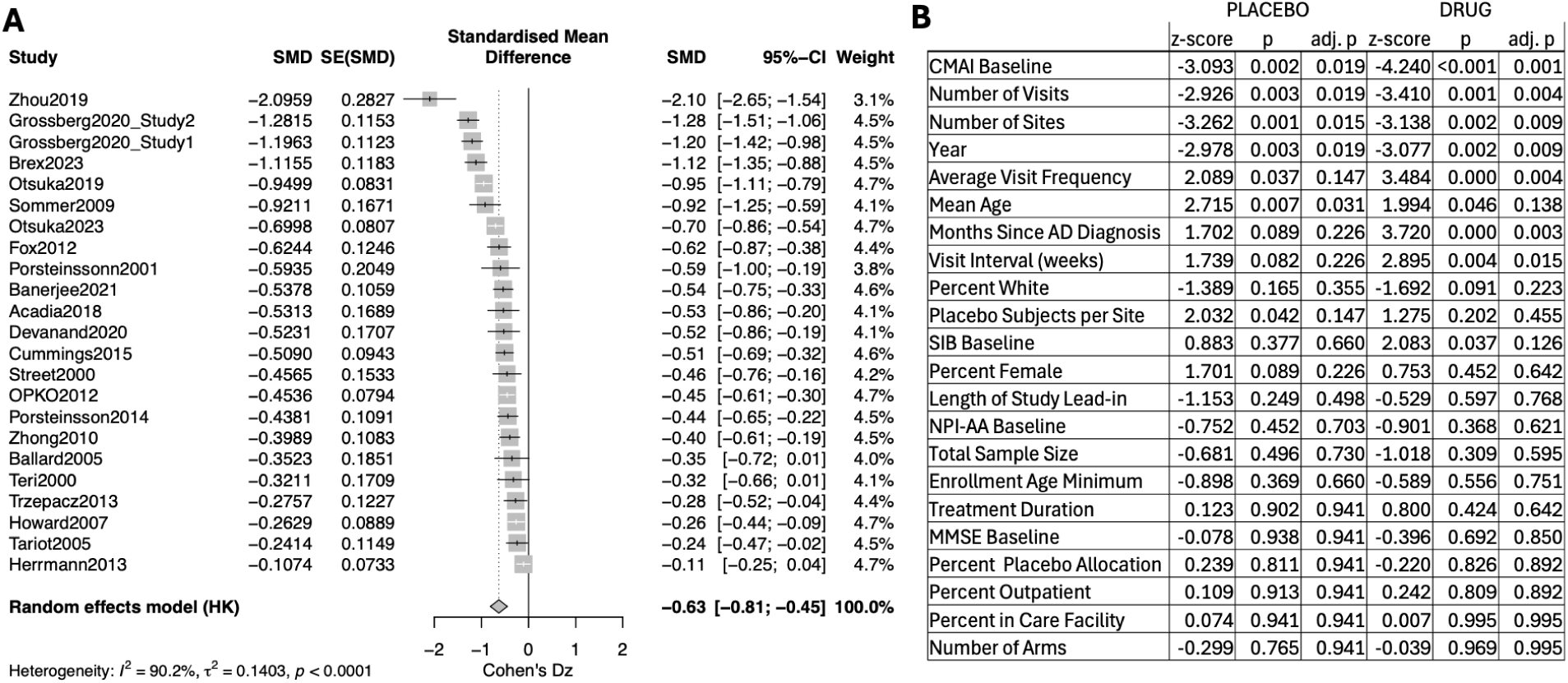
Study-level predictors of placebo response tend to also predict drug response. A) Standardized effect size (Cohen’s d_z_) values for placebo arm change from baseline in clinical scale (CMAI or NPI-AA). B) Metaregression results for association between study-level factors and change from baseline in drug and placebo arms. SMD=Standardized Mean Difference; SE=Standard Error; HK=Confidence interval method by Hartung and Knapp. Black or white bars show 95% confidence interval.

### Reducing placebo response in clinical trials of agitation in Alzheimer’s Disease using causal modeling

We then moved to analyses of individual patients, aimed at increasing the resolution to distinguish drug from placebo effects and used causal modeling to estimate the impact of clinical characteristics on treatment/placebo response of an atypical and a typical antipsychotic (risperidone and haloperidol) for agitation in Alzheimer’s disease (AD). These classes of models leverage counterfactual states to make inferences about a patient’s predicted change in an endpoint when given the active drug and when given placebo, and their expected difference, termed the “conditional average treatment effect” (CATE).

Unlike analyses of subjects only receiving placebo – akin to the meta-regression analyses above, or analyses of a single placebo arm ^38^– these causal models can help distinguish between factors that predict increased response in either arm to those specific to drug or placebo.

We used historical clinical trial datasets of risperidone in AD patient populations that measured improvements in the Cohen Manfield Agitation Inventory (CMAI) ^25^ as a key endpoint (see Methods). We use two related criteria for evaluating model performance: a) inference at the individual patient level, corresponding to the accuracy (coefficient of determination R^2^) of predicted CMAI change in the observed/factual treatment, and b) inference at the study level, with high separability in treatment response after stratifying by (baseline) CATE scores, e.g. comparing patients predicted to be more drug responsive (bottom 75%) versus more placebo responsive (top 25%).

We fit a linear X-Learner causal model to the training dataset (RIS-AUS-5 clinical trial, N=305), predicting week 12 change in CMAI at the trial conclusion as a function of baseline clinical symptomatology and demographic data (see Methods). This model takes subject-level baseline covariates/variables as input and produces (a) the predicted CMAI change when on drug, (b) the predicted CMAI change when on placebo and (c) a prediction of their difference—CATE— for each subject. Within the training dataset, we compared model-predicted changes in CMAI against observed changes in CMAI under actual/observed treatment arms at the individual/patient level, and found significant accuracy both overall (R^2 = 0.31, p=3.0e-28), and within each treatment arm (placebo arm R^2^= 0.39, p = 6.7e-19; risperidone arm R^2^ = 0.21, p = 1.7e-10; Figure 2A).

**Figure 2:**
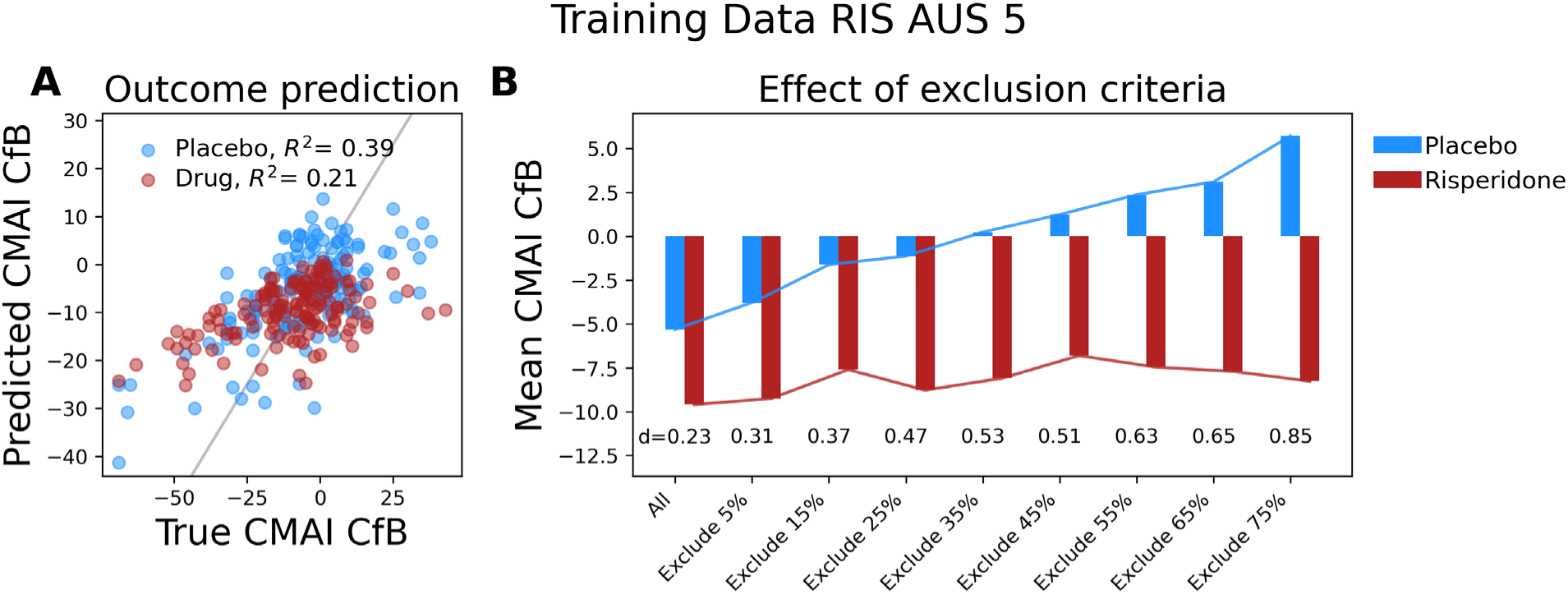
Exclusion of placebo-indicated subjects increases treatment effects. Training results: A) Individuals’ true vs. predicted change in CMAI in each arm of the training RIS-AUS-5 dataset and B) the study-level impact of dropping high-CATE participants on the mean CMAI improvement in the placebo and drug arms of the training RIS-AUS-5 dataset (bottom annotations show Cohen’s d).

We then contrasted the efficacy of risperidone versus placebo in our full set of participants (N=305) before and after stratifying by outputs of this causal model, specifically within drug-indicated (bottom 75% of CATE, ≤ -1.7, N = 228) and placebo-indicated (top 25% of CATE, > -1.7 N = 77 ) subgroups. In the full set of 305 participants, the average CMAI change from baseline was –9.6 in the risperidone arm, and -5.3 in the placebo arm, with a placebo-adjusted treatment effect of –3.9 (p=0.03, via ANCOVA for the change in CMAI, controlling for baseline CMAI). Excluding the placebo-indicated (CATE > -1.7) subgroup increased the placebo-adjusted treatment effect to –7.6 (p=0.0001), with an average CMAI change of –8.8 in the risperidone arm and – 1.1 in the placebo arm. There were large decreases in CMAI among both treatment arms within this placebo-indicated subgroup (risperidone = -11.7, placebo = -19.3) and in the placebo-indicated subgroup the placebo-and baseline-severity adjusted treatment effect numerically favored placebo (+7.9, p=0.06). Overall, therefore, placebo-response causal model showed high performance in the training data at the levels of individual patients and overall trial efficacy.

### Strong replication of the placebo response model in an independent dataset

Prediction models built in training datasets may be over-fit and not generalize well to independent testing datasets. For instance, models fit on one clinical trial may have poor predictive accuracy when applied prospectively to another trial. Therefore, we evaluated the generalizability and replicability of our final “CMAI Placebo Response Model” by applying it to the independent/unseen RIS-INT-24 dataset. We generated a CATE score for each subject in RIS-INT-24 data by applying the final causal model to baseline covariates across each of the N=302 subjects with complete baseline data, of which 100 were randomized to placebo, 97 randomized to risperidone, and 105 randomized to haloperidol, a typical antipsychotic medication not present in the training dataset.

We first replicated the ability for this model to predict changes in CMAI at the individual patient level within the placebo (R^2 = 0.24, p= 3.3e-6) and risperidone (R^2 = 0.19, p = 3.5e-6) treatment arms (Figure 3A). Haloperidol was not present in the training dataset, and the accuracy of predicting the haloperidol-randomized participants with the risperidone model (R^2 = 0.085, p = 0.003) or placebo model (R^2 =0.083, p = 0.003) were nearly identical.

**Figure 3:**
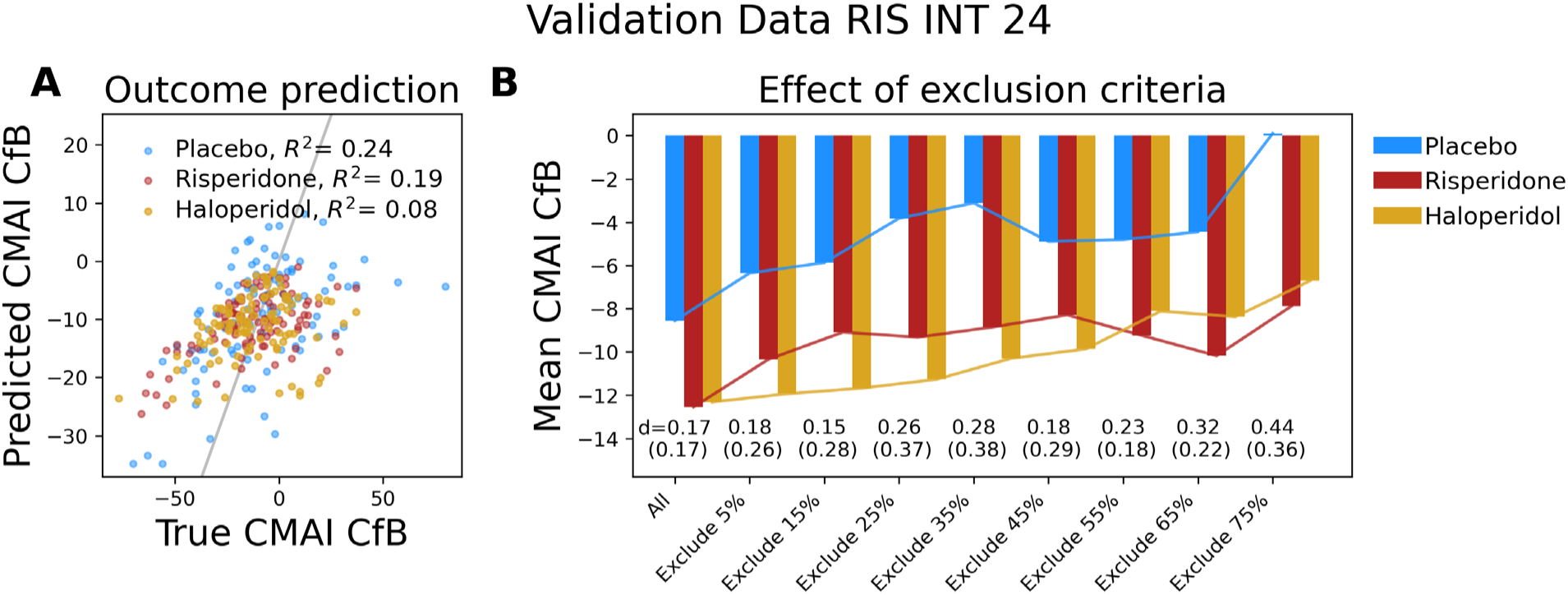
External validation replicates pattern of increased treatment effects through filtering of placebo indicated patients. A) True vs. Predicted change in CMAI in each arm of the validation RIS-INT-24 dataset. The performance in the validation dataset is not far below training performance, indicating generalizability. B) CMAI change scores in the validation set, excluding participants with the highest (most placebo-favorable) CATE estimates at baseline. Percentile CATE thresholds chosen according to the training dataset CATE percentiles. Annotations show the Cohen’s d effect size for risperidone (haloperidol). As participants are excluded, the improvement attenuates in both arms, but the placebo response decreases more than the treatment response, resulting in generally larger difference between treatments and placebo and larger effect size.

We then sought to replicate the study-level enrichment of placebo-adjusted treatment efficacy. That is, we assessed whether we could increase the probability of trial success by prospectively assigning each patient to placebo- or drug-indicated subgroups based on their baseline-derived CATE score and using a threshold of CATE > -1.7 (the cutoff designated in the training dataset). In the full set of validation data, the average CMAI change from baseline was -12.5 in the risperidone arm, (n=197), -12.3 in the haloperidol arm (n=197), and –8.6 in the placebo arm (n=100) with placebo- and baseline- adjusted treatment effects of -5.3 or risperidone (p=0.048), and -4.3 for haloperidol (p=0.12) and a combined/joint drug treatment effect of –4.7 (p=0.045, via ANCOVA controlling for baseline CMAI). Among the drug-indicated subgroup (CATE ≤-1.7, N=229), while the average CMAI change from baseline attenuated in each active arm (risperidone arm (-9.3, n=72) and haloperidol arm (-11.3, n=78)), it greatly attenuated in the placebo arm (-3.8, n=72), producing placebo- and baseline-adjusted treatment effects that were stronger and – despite the reduced arm size— more significant for risperidone (-6.5, p = 0.046), haloperidol (-7.8, p = 0.01) and the drug arms combined (-7.1, p=0.007). Within the placebo-indicated subgroup (CATE > -1.7, N=80), there were again large decreases in CMAI in all three treatment arms (risperidone = -21.8 haloperidol = -15.3 placebo =-20.8) similar to what was observed in the training dataset, and placebo-adjusted treatment effects for each drug that numerically favored placebo as compared to haloperidol (haloperidol: +5.3, p=0.4,) and both drug arms combined (+1.9, p=0.7) , and was greatly attenuated for risperidone ( -2.0 , p=0.7). . These study-level enrichments were highly comparable with (and perhaps stronger than) those calculated in the training dataset, a rare feat in machine learning prediction modeling.

We performed several sensitivity analyses around aspects of the placebo response model building and its application. First, we evaluated the CATE threshold used to designate “placebo-indicated” or “drug-indicated” subgroups at baseline in the validation dataset by using different thresholds (e.g. less than or greater than –1.7) and evaluated the subsequent treatment efficacy in the drug-indicated subgroup. Larger exclusions of placebo-indicated subjects generally produced larger placebo-adjusted treatment effects, with larger attenuations of CMAI changes from baseline in the placebo arm (Figure 2B, 3B, S4B)

**Table 1:**
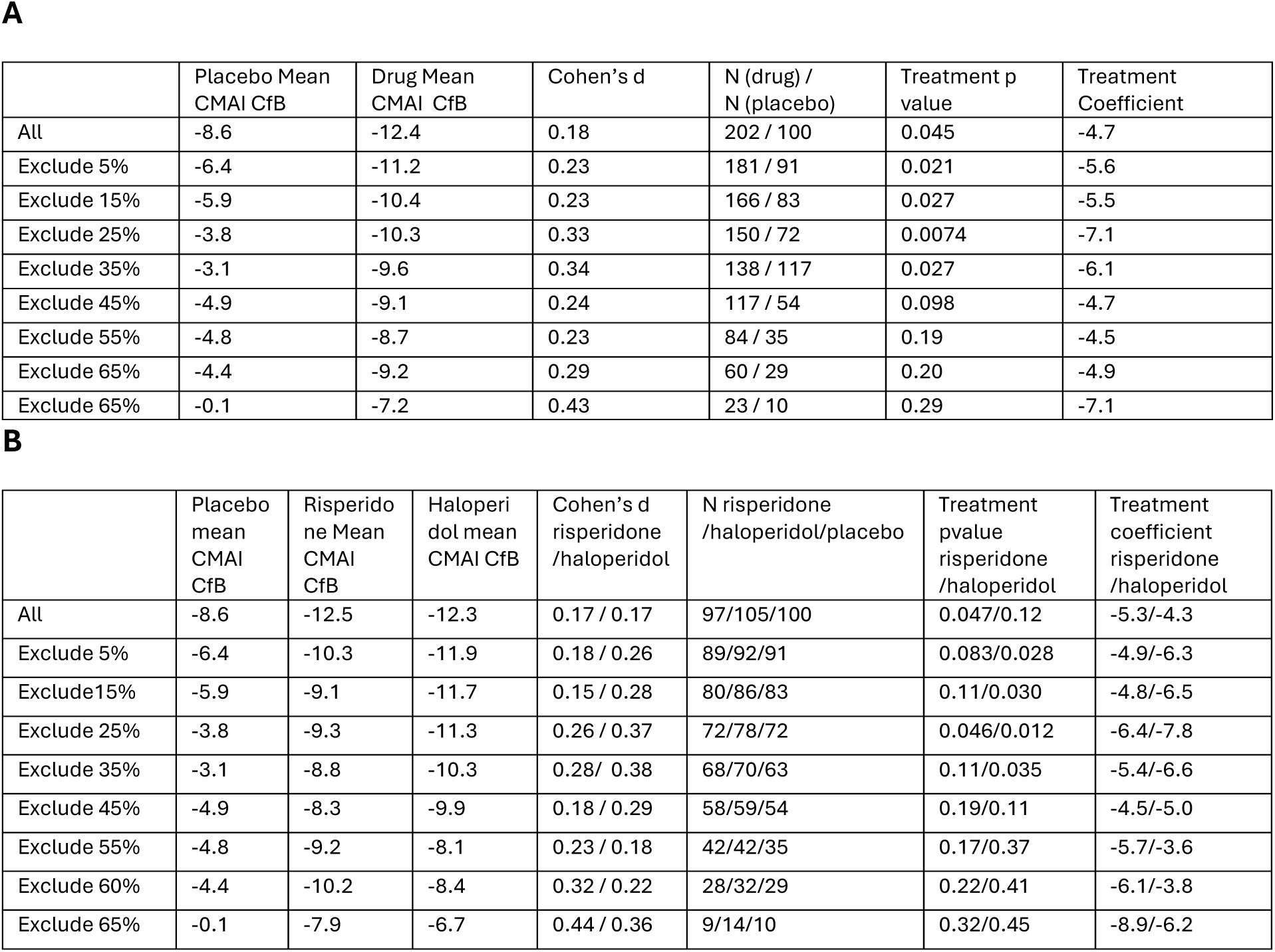
A comparison of estimates of treatment effect and associated significance estimated with an ANCOVA controlling for baseline severity, as participants are excluded from the validation set based on baseline model-produced CATE estimates and percentile cutoffs from the training data. In Table A, the two drug arms are combined. In Table B, they are each separately compared to placebo. CfB=Change from baseline

|  | Placebo Mean<br>CMAI CfB | Drug Mean<br>CMAI CfB | Cohen's d | N (drug) /<br>N (placebo) | Treatment p<br>value | Treatment<br>Coefficient |
| --- | --- | --- | --- | --- | --- | --- |
| All | -8.6 | -12.4 | 0.18 | 202 / 100 | 0.045 | -4.7 |
| Exclude 5% | -6.4 | -11.2 | 0.23 | 181 / 91 | 0.021 | -5.6 |
| Exclude 15% | -5.9 | -10.4 | 0.23 | 166 / 83 | 0.027 | -5.5 |
| Exclude 25% | -3.8 | -10.3 | 0.33 | 150 / 72 | 0.0074 | -7.1 |
| Exclude 35% | -3.1 | -9.6 | 0.34 | 138 / 117 | 0.027 | -6.1 |
| Exclude 45% | -4.9 | -9.1 | 0.24 | 117 / 54 | 0.098 | -4.7 |
| Exclude 55% | -4.8 | -8.7 | 0.23 | 84 / 35 | 0.19 | -4.5 |
| Exclude 65% | -4.4 | -9.2 | 0.29 | 60 / 29 | 0.20 | -4.9 |
| Exclude 65% | -0.1 | -7.2 | 0.43 | 23 / 10 | 0.29 | -7.1 |

B
|  | Placebo<br>mean<br>CMAI<br>CfB | Risperido<br>ne Mean<br>CMAI<br>CfB | Haloperi<br>dol mean<br>CMAI CfB | Cohen's d<br>risperidone<br>/haloperidol | N risperidone<br>/haloperidol/placebo | Treatment<br>pvalue<br>risperidone<br>/haloperidol | Treatment<br>coefficient<br>risperidone<br>/haloperidol |
| --- | --- | --- | --- | --- | --- | --- | --- |
| All | -8.6 | -12.5 | -12.3 | 0.17 / 0.17 | 97/105/100 | 0.047/0.12 | -5.3/-4.3 |
| Exclude 5% | -6.4 | -10.3 | -11.9 | 0.18 / 0.26 | 89/92/91 | 0.083/0.028 | -4.9/-6.3 |
| Exclude 15% | -5.9 | -9.1 | -11.7 | 0.15 / 0.28 | 80/86/83 | 0.11/0.030 | -4.8/-6.5 |
| Exclude 25% | -3.8 | -9.3 | -11.3 | 0.26 / 0.37 | 72/78/72 | 0.046/0.012 | -6.4/-7.8 |
| Exclude 35% | -3.1 | -8.8 | -10.3 | 0.28 / 0.38 | 68/70/63 | 0.11/0.035 | -5.4/-6.6 |
| Exclude 45% | -4.9 | -8.3 | -9.9 | 0.18 / 0.29 | 58/59/54 | 0.19/0.11 | -4.5/-5.0 |
| Exclude 55% | -4.8 | -9.2 | -8.1 | 0.23 / 0.18 | 42/42/35 | 0.17/0.37 | -5.7/-3.6 |
| Exclude 60% | -4.4 | -10.2 | -8.4 | 0.32 / 0.22 | 28/32/29 | 0.22/0.41 | -6.1/-3.8 |
| Exclude 65% | -0.1 | -7.9 | -6.7 | 0.44 / 0.36 | 9/14/10 | 0.32/0.45 | -8.9/-6.2 |

### Clinical characteristics of placebo-indicated AD patients

Given the similarly high model performance in both the training and testing datasets, we next examined clinical characteristics of the placebo response model itself. We first correlated the model-produced CATE scores with each baseline characteristic to determine the marginal/univariate contributions in the RIS-AUS-5 dataset (Figure 4).

**Figure 4:**
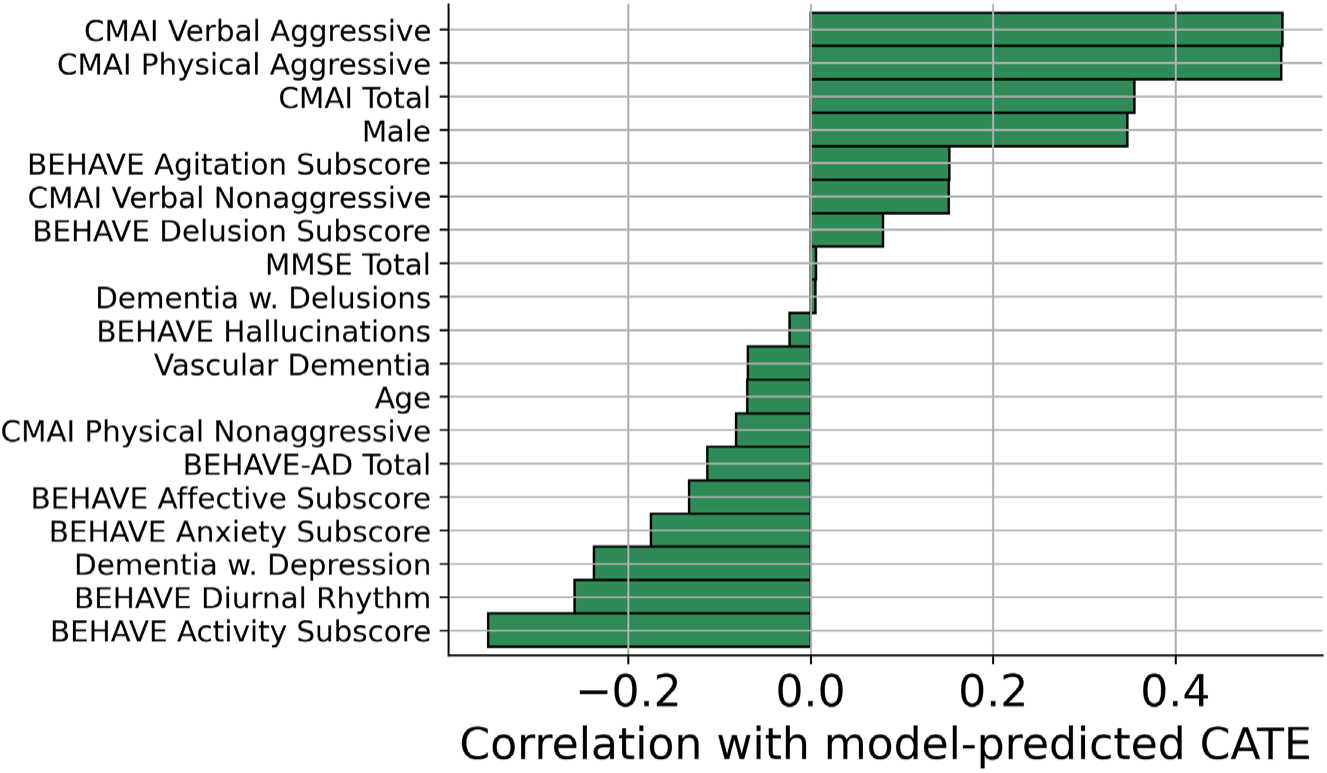
Higher baseline CMAI and CMAI aggressive subscores correlate with higher estimated placebo response relative to drug response. Univariate correlations of each baseline covariate with CATE estimates in the RIS-AUS-5 training dataset. Positive correlations mean that higher values of the covariate associate with better placebo response relative to drug response, and negative correlations mean that higher values of the covariate associate with better drug response relative to placebo response.

Severity of verbal and physical aggression and overall agitation as measured by the CMAI total score were most highly correlated with increased predicted placebo response relative to risperidone response (i.e. more positive CATE values) whereas presence of BEHAVE-AD-measured activity disturbances, diurnal disturbances, and affective, anxiety or depressive symptoms were more associated with increased predicted risperidone treatment response relative to placebo response.

We therefore stratified AD patients by their baseline CMAI severity and examined the clinical characteristics of the resulting subgroups. The most severe patients (CMAI > 100) had much more prevalent physical aggression, particularly on items related to hurting self or others, pushing, kicking and scratching, and aspects of verbal aggression like screaming (Figure S6). For instance, participants with CMAI above 100 were described as hurting themselves or others on average “Once or twice a day”, compared to an average score of “Less than once a week” among all participants with CMAI above 45.

We lastly assessed the potential for using CMAI total score and aggressive subscores as surrogate for the more complex placebo response model in attenuating placebo response at the study level. We evaluated a series of thresholds/caps of CMAI total score and the physical and verbal aggressive subscores as inclusion criteria in the combined RIS-AUS-5 and RIS-INT-24 datasets. We found that excluding more severe patients attenuated the CMAI change from baseline in both the active and placebo arms, with larger effects in the placebo arms, resulting in larger placebo-adjusted risperidone effects (Cohen’s d) (Figure 5). For instance, filtering who were above the 75^th^ percentile of CMAI led to an increase of Cohen’s d from 0.23 to 0.37.

**Figure 5:**
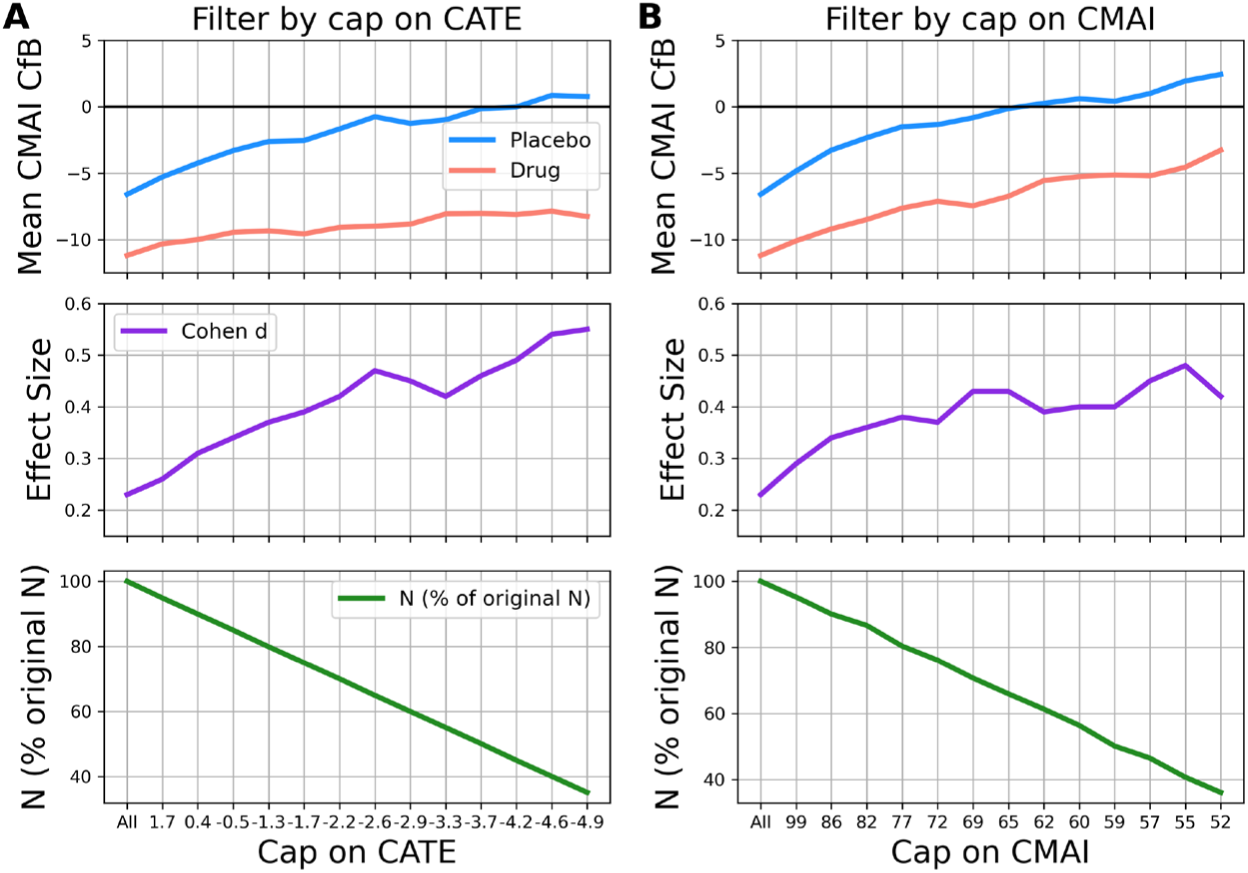
Simple thresholding of model-generated CATE values or baseline CMAI leads to increased treatment effect. The effect within the combined training and validation datasets of baseline cap of CATE (A) and CMAI (B) on CMAI change scores in each arm (top), Cohen’s d effect size (middle) and percentage of the original participants who remained after the cap (bottom). The bottom panels allow for assessing the tradeoffs in terms of fraction of patients excluded that correspond to the increase in effect size shown in the middle pane.

### Generalizing the placebo response model in trials of acetylcholinesterase inhibitors

We demonstrated generalizability of our placebo response model across training and testing datasets of clinical trials of antipsychotics, and the contribution of symptom severity as a contributing factor. However, both trials were highly similar and evaluated the same therapeutics and measured the same clinical scales, such that we could not fully determine if placebo-indicated subgroup captures those that would poorly respond to risperidone rather than better respond to placebo. For example, we saw lower and non-differential performance at the subject level applying the final causal model to patients treated with haloperidol, but similar improvements in trial-level efficacy. We therefore evaluated further generalizability using two clinical trials of galantamine, an acetylcholinesterase inhibitor originally aimed at improving cognitive endpoints in AD (GAL-USA-10 and GAL-INT-10, see Methods).

Both galantamine trials used the Neuropsychiatric Inventory (NPI) scale for characterizing BPSD, and did not include CMAI. Therefore, we evaluated our placebo response model using the NPI Agitation/Aggression (NPI-AA) subscale as the CMAI surrogate for agitation severity at baseline and as an outcome. We included in our analysis those participants with clinically significant agitation/aggression at baseline (NPI-AA ≥ 4, N=220), and combined the two studies into a single dataset large enough to enable subgroup analyses.

The most severely agitated patients in these studies of galantamine again had larger placebo responses in agitation—both in absolute terms and relative to the drug response of patients with the same severity—and excluding patients based on symptom severity both attenuated placebo response and increased the placebo-adjusted treatment effects (Figure 6, Figure S7). As the baseline severity was capped with increasing strictness, the size and significance of the treatment effect increased (Table 3). These results suggest the interplay between increased baseline symptom severity and subsequent increased placebo response generalizes across drug mechanisms.

**Figure 6:**
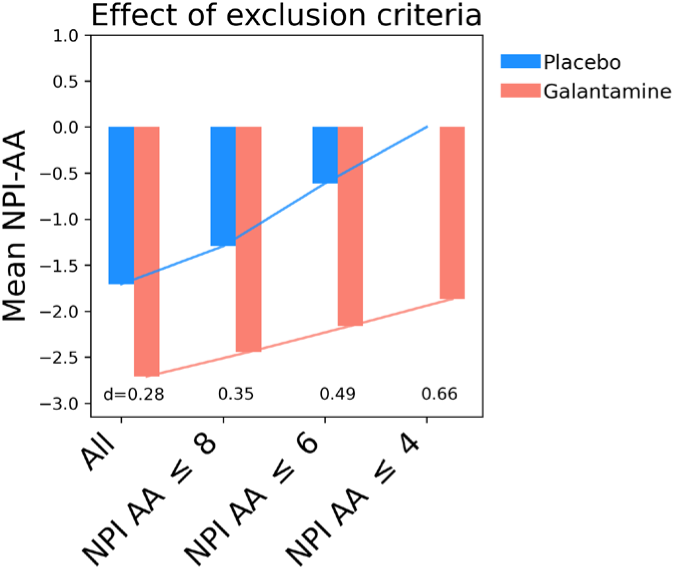
A cap on baseline NPI-AA severity within agitated subset of a participants increases the effect size in the NPI-AA outcome in pooled studies of galantamine.. Compared to including all participants with clinically significant agitation at baseline (“All”), adding an upper limit on baseline severity of 8, 6, or 4, decreased the placebo response in absolute terms and relative to drug response. Annotations are Cohen’s d effect sizes.

**Table 3:** Effect of setting a cap on baseline NPI-AA severity on NPI-AA agitation outcome in the agitated subset of the pooled Galantamine study participants. Treatment effects and associated are estimated using an ANCOVA on NPI-AA change score, controlling for baseline severity.

| Range of NPI-AA | N total (pbo/drug) | Treatment effect | p-value |
| --- | --- | --- | --- |
| $\geq 4$ | 220 (65/155) | -1.4 | 0.14 |
| Between 4 and 8 | 196 (61/135) | -2.0 | 0.046 |
| Between 4 and 6 | 155 (49/106) | -2.6 | 0.010 |
| = 4 | 84 (31/53) | -2.9 | 0.004 |

## DISCUSSION

In this study, we demonstrate that causal models can increase the likelihood of clinical trial success through the reduction of placebo response. High placebo response remains a key barrier to successful development of treatments for AAD, potentially obscuring true drug effects and leading to costly clinical trial failures. Using historical trials of risperidone and haloperidol, we demonstrate that a causal model trained on baseline clinical features generalizes across independent studies, and that prospectively excluding patients predicted to have high placebo response markedly increases placebo-adjusted treatment effects.

The meta-analysis and metaregression results allowed for characterization of the magnitude of the placebo response in trials for BPSD and identification of trial-level features (number of sites, number of visits and mean baseline severity, year of study and mean age) that associated with placebo response. The analysis also served to underscore a key challenge in extracting actionable insights about placebo response from summary data. Since all but one (mean age) of the predictors significantly associated with placebo response related to drug response as well, these study-level factors do not easily lend themselves to forming a strategy that would reduce placebo response while preserving drug response.

Therefore, in the second part of the analysis, we used subject-level data from clinical trials of risperidone to build and then independently validate a placebo response model of agitation using causal inference methodologies. We predicted placebo response with meaningful accuracy at the level of individual patients (e.g. accurate predictions of outcome improvements when randomized to placebo, R^2^ =0.24 in validation set) and overall trials (e.g. increased/enriched treatment effect when removing predicted placebo-responsive subjects). Primary components of this placebo response model strongly related to underlying symptom severity underlying symptom severity within the domain of agitation and aggression, a relationship which appeared to generalize to the treatment effects of galantamine on neuropsychiatric symptoms of AD. Moreover, we observed that the predicted placebo responders had a high level of improvement regardless of their actual treatment arm. These results support the idea that individuals with the highest baseline agitation scores do not have as much relative benefit (compared to the benefit of placebo), and that excluding them could increase placebo-adjusted efficacy.

Generalizability is crucial to the applicability of clinical trial modelling to drug development but achieving it has proved challenging^39^. We therefore assessed both individual-level and study-level generalizability and demonstrated the presence of both success criteria. While most research in the area focuses on the first metric—accurate prediction of individual-level outcomes—as the standard for model success^39^, we would argue that it is the second, study-level, criterion that is more directly related to a model’s potential impact on future clinical trials. The study-level metrics allow for measuring the impact on trial-level efficacy measures of strategies like prospectively excluding the most placebo-responsive patients. Moreover, they allow for assessment of the tradeoffs, e.g. between the downsides of a potential inclusion/exclusion criterion (e.g. overall study enrollment slowdowns associated with any narrowing of eligibility) and possible increases in efficacy measures (e.g. as highlighted in Figure 5).

Accurate prospective prediction—and subsequent exclusion— of placebo-responsive patients could greatly increase the probability of technical and regulatory success (PTRS) of a therapeutic agent. Moreover, additional strategies exist. As an alternative to an exclusion-based strategy, especially in earlier stages or where less is known, modeling results could be used to inform the selection of prespecified subgroups for analysis, so that information about the relative responsiveness of such groups to drug or placebo response can be assessed and evidence accumulated over time.

We found that excluding participants at baseline using the model-derived placebo response predictions—even in unseen datasets with different drug mechanisms—enhanced placebo-adjusted treatment effects, and often statistical significance (even though the resulting patient population was smaller). Interestingly, even though the individual-level predictions for haloperidol were less accurate than those for risperidone, the study-level exclusion strategy was more effective at increasing the placebo-adjusted treatment effect of haloperidol compared to risperidone, a result that highlights the importance of considering the trial-level generalizability results alongside patient level predictive accuracy.

These results also serve to underscore the advantages of causal modeling methodologies, which make use of data from patients that received drug and placebo, for identifying factors associated with placebo response. Previous work has analyzed a single placebo arm and found that higher baseline scores predict greater improvement on placebo, but without the causal approach or an assessment of how these same scores relate to treatment response, such results do not by themselves give information about the on study results or the difference between drug and placebo arms^38^. Our causal modeling work adds the key information that in the studies we considered at the patient-level, baseline agitation severity predicted not only placebo response but also a more placebo-favorable difference between drug and placebo response.

To explain the importance of these causal predictions more generally, predicting response within subjects that only received placebo (e.g. in the placebo arm) could result in both false positive and false negative predictors. If the same factors associated with placebo improvement also associate with drug improvement, then eliminating placebo-responders using this strategy may also exclude those who would benefit most from the drug. It is not enough to predict responses within a placebo arm and identify factors that relate to higher or lower placebo response; instead, we need to understand how baseline factors relate to how much more beneficial the drug treatment will be for an individual than the placebo treatment. A causal modelling approach allows for individual-level baseline estimation of the difference between the potential outcomes on drug vs placebo, which can then be used to include or exclude participants or make other adjustments that impact study-level results.

Although the strong transferability of the results from one trial to another among those considered is a promising result, we note that these were all trials from the same sponsor, conducted within a similar time frame. We were able to validate the relationship of baseline severity to drug-placebo separation in the agitated subpopulation of the galantamine trials, but could not validate the full model due to differences in the scales used to measure BPSD. Validating the model in more recent AAD trials, trials by a different sponsor, or trials of a wider variety of drugs would give important information about its broader applicability.

## CONCLUSION

We assessed predictors of placebo response in agitation in Alzheimer’s disease through metaregression and causal modeling. Here the causal modelling approach showed strong generalizability to an independent dataset, according to both individual and—key to its practical importance— trial-level measures. This application of causal modelling to clinical trials for agitation in Alzheimer’s disease gives promising directions for strategies to identify participants during screening (or prior to randomization) predicted to have especially high placebo response, and vice versa. For instance, participants with more placebo-indicated pre-treatment CATE values could be excluded. Alternatively, the one-variable proxies for the full-model CATE could be used as an inclusion/exclusion criterion incorporated into trial design to enroll participants who are more likely to have a greater benefit from the drug relative to placebo. The trial-level metrics used here are designed to assess the potential effectiveness of such a strategy, and are an important complement to more common metrics that assess the reliability of model predictions at the individual level. Given the recent and rapid expansion of causal machine learning methodologies, their applicability to clinical trials is likely to continue to expand, and we hope that future work can continue to uncover predictors—including those specific to different drug mechanisms—that separate drug and placebo response, and can be leveraged to minimize the degree to which excessive placebo response hinders accurate assessment of drug efficacy and to increase the PTRS of effective new therapeutics.

## Data Availability

This study, carried out under YODA Project # 2023-5295, used data obtained from the Yale University Open Data Access Project, which has an agreement with JANSSEN RESEARCH & DEVELOPMENT, L.L.C.. The interpretation and reporting of research using this data are solely the responsibility of the authors and does not necessarily represent the official views of the Yale University Open Data Access Project or JANSSEN RESEARCH & DEVELOPMENT, L.L.C..

https://yoda.yale.edu/

## Acknowledgements

This study, carried out under YODA Project # 2023-5295, used data obtained from the Yale University Open Data Access Project, which has an agreement with JANSSEN RESEARCH & DEVELOPMENT, L.L.C.. The interpretation and reporting of research using this data are solely the responsibility of the authors and does not necessarily represent the official views of the Yale University Open Data Access Project or JANSSEN RESEARCH & DEVELOPMENT, L.L.C.. K.C.K., M.B., N.J.B. and A.E.J. are current employees and shareholders of Neumora Therapeutics. D.S. is a current consultant and shareholder of Neumora Therapeutics. K.M.A., R.A.L., T.D., and T.B. are former employees and potential shareholders of Neumora Therapeutics (and were employees of Neumora Therapeutics when the work in this manuscript was performed).

## Supplementary material

**Figure S1:**
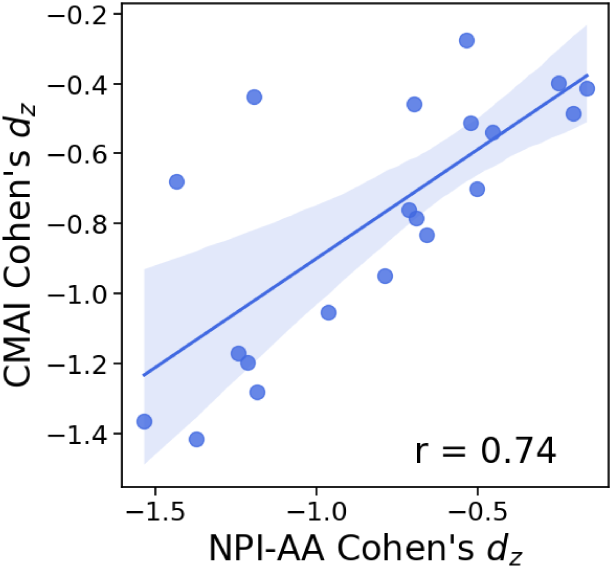
CMAI and NPI-AA standardized change scores are highly correlated. For study arms that measured both CMAI and NPI-AA, the correlation between the standardized change scores across these scales was 0.74 (or 0.93 after dropping the two Porsteinsson2014 arms, which appear as outliers in the upper right portion of the plot).

**Figure S2:**
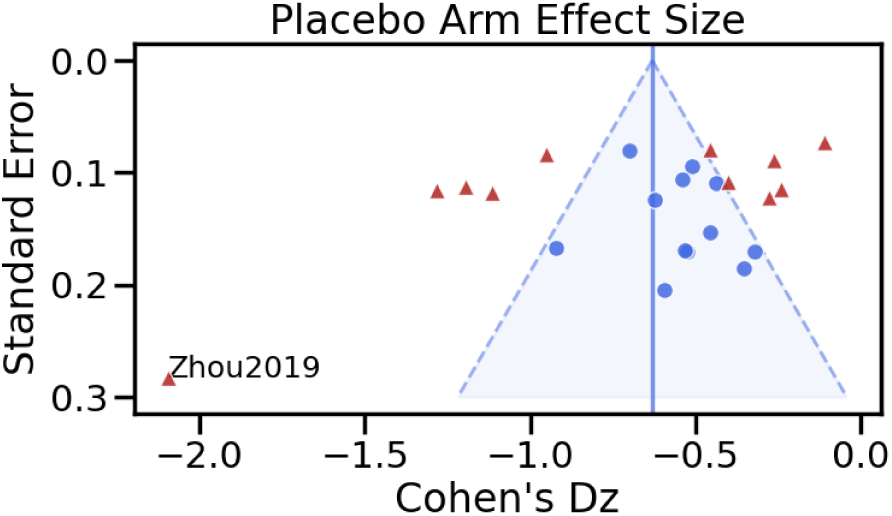
Funnel plot for placebo arm effect sizes shows possible asymmetry towards smaller placebo effect sizes.

**Figure S3:**
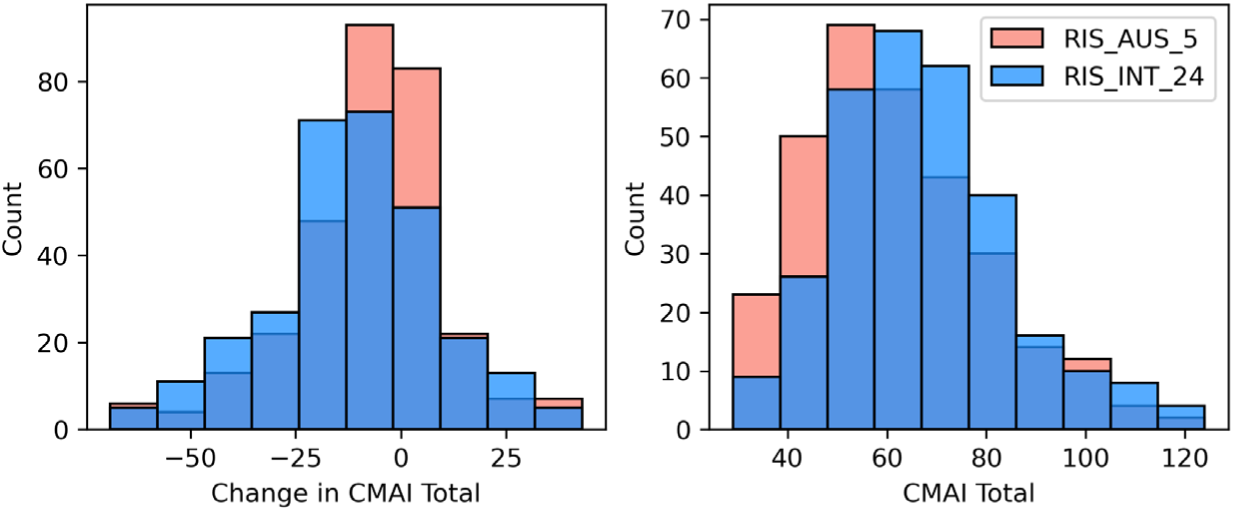
Distribution of change in CMAI and baseline CMAI severity are comparable in training and validation datasets.

**Figure S4:**
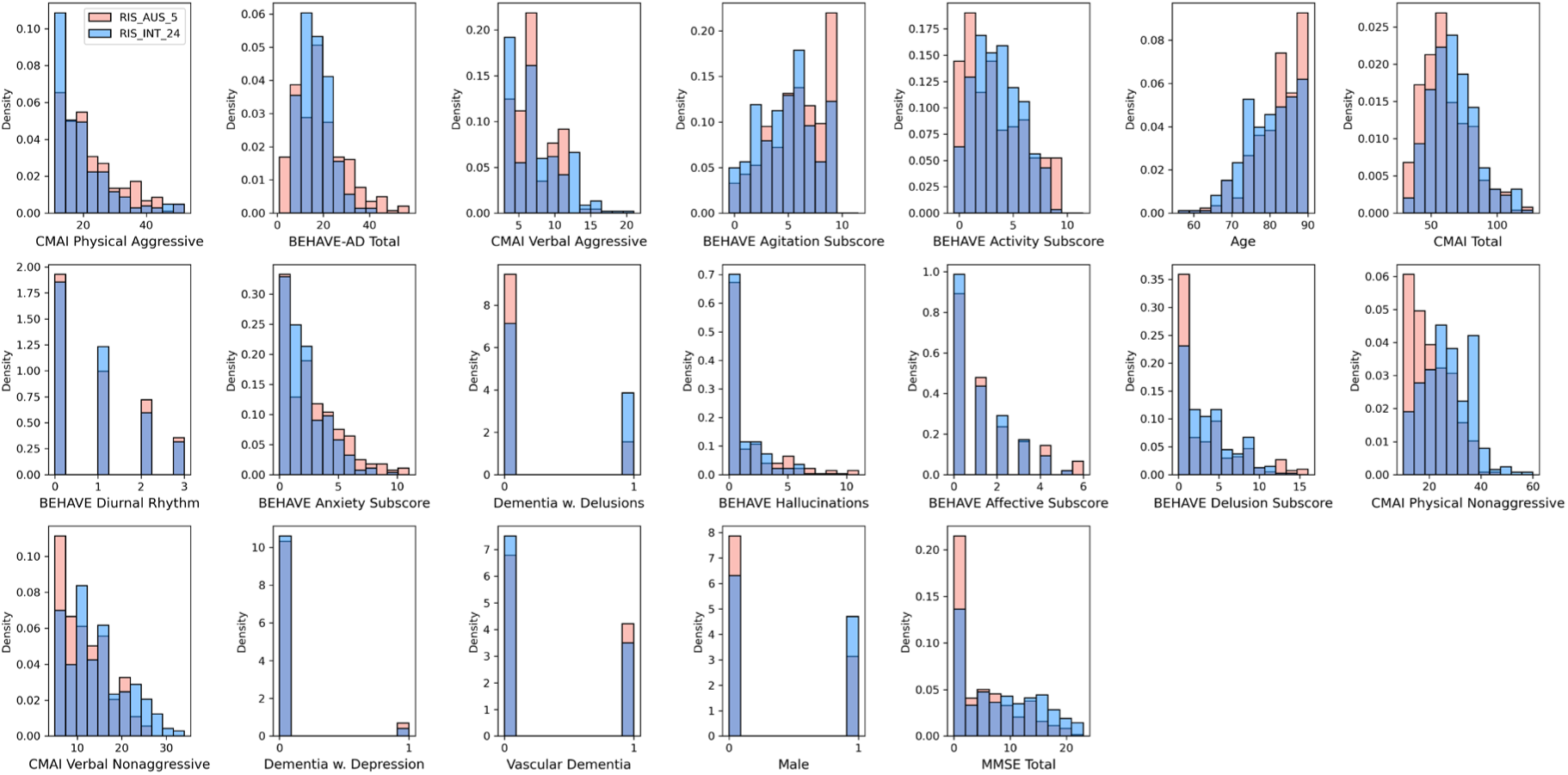
Participants in the training and validation set have similar baseline characteristics.

**Figure S5.**
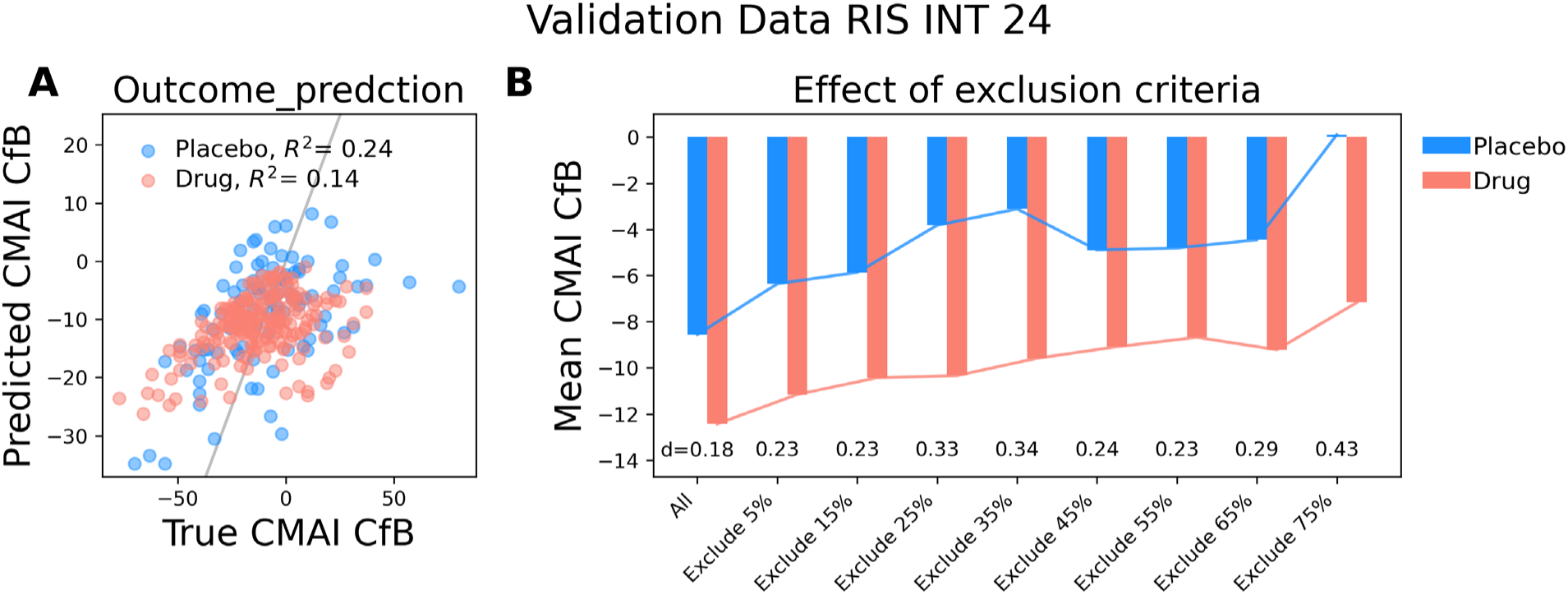
Excluding patients by estimated CATE score in the validation set generally increases the effect size also in the case where the risperidone and haloperidol arms are pooled. Figure is as in Figure 3, but with risperodine and haloperidol drug arms combined.

**Figure S6:**
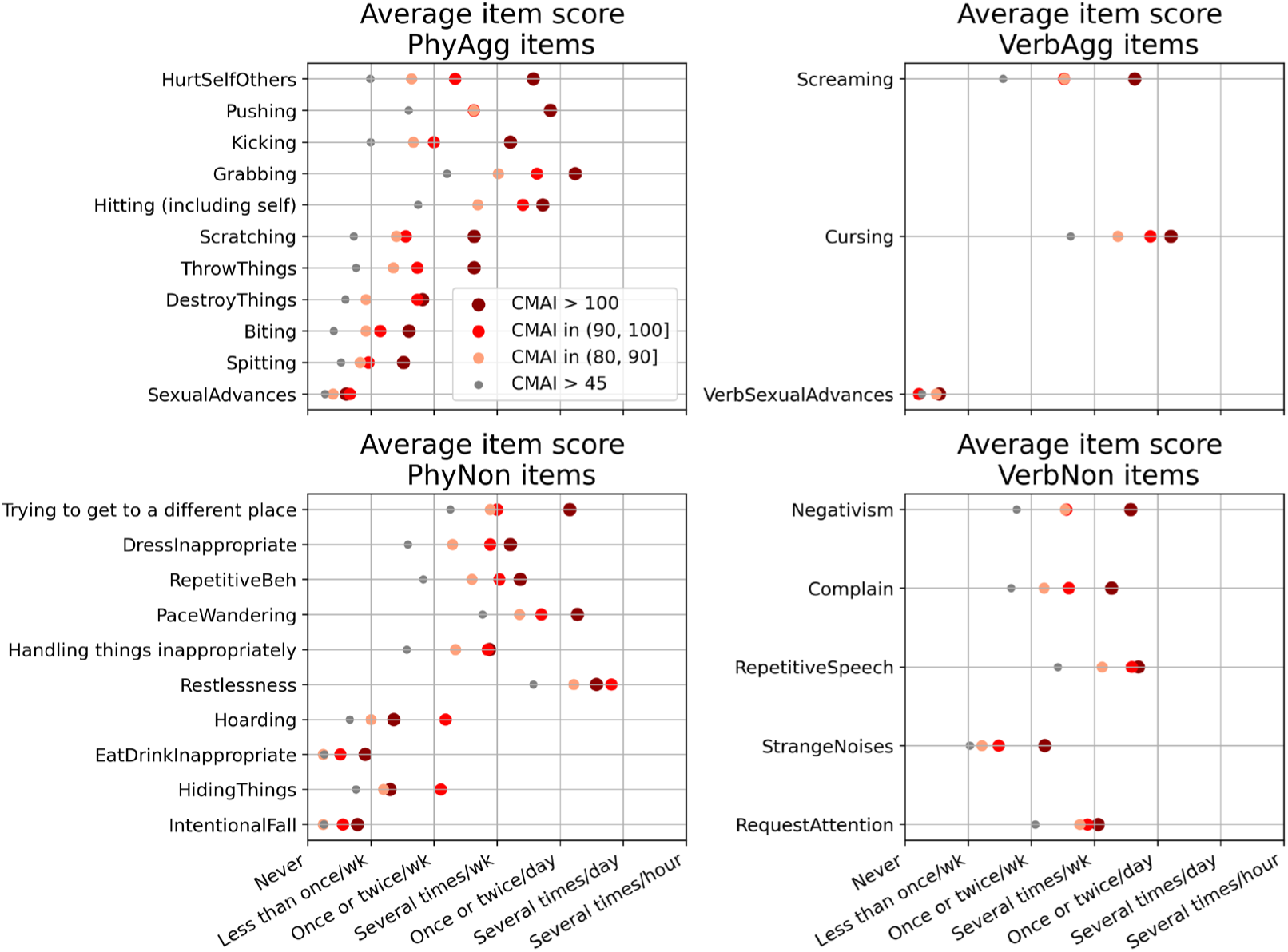
The CMAI scale is based on frequency measures of behaviors that include physical and verbal as well as aggressive and non-aggressive domains. Frequency of CMAI-measured behaviors observed in three CMAI severity bands (>100, (90,100], (80,90], compared to those in all participants with total CMAI > 45.

**Figure S7:**
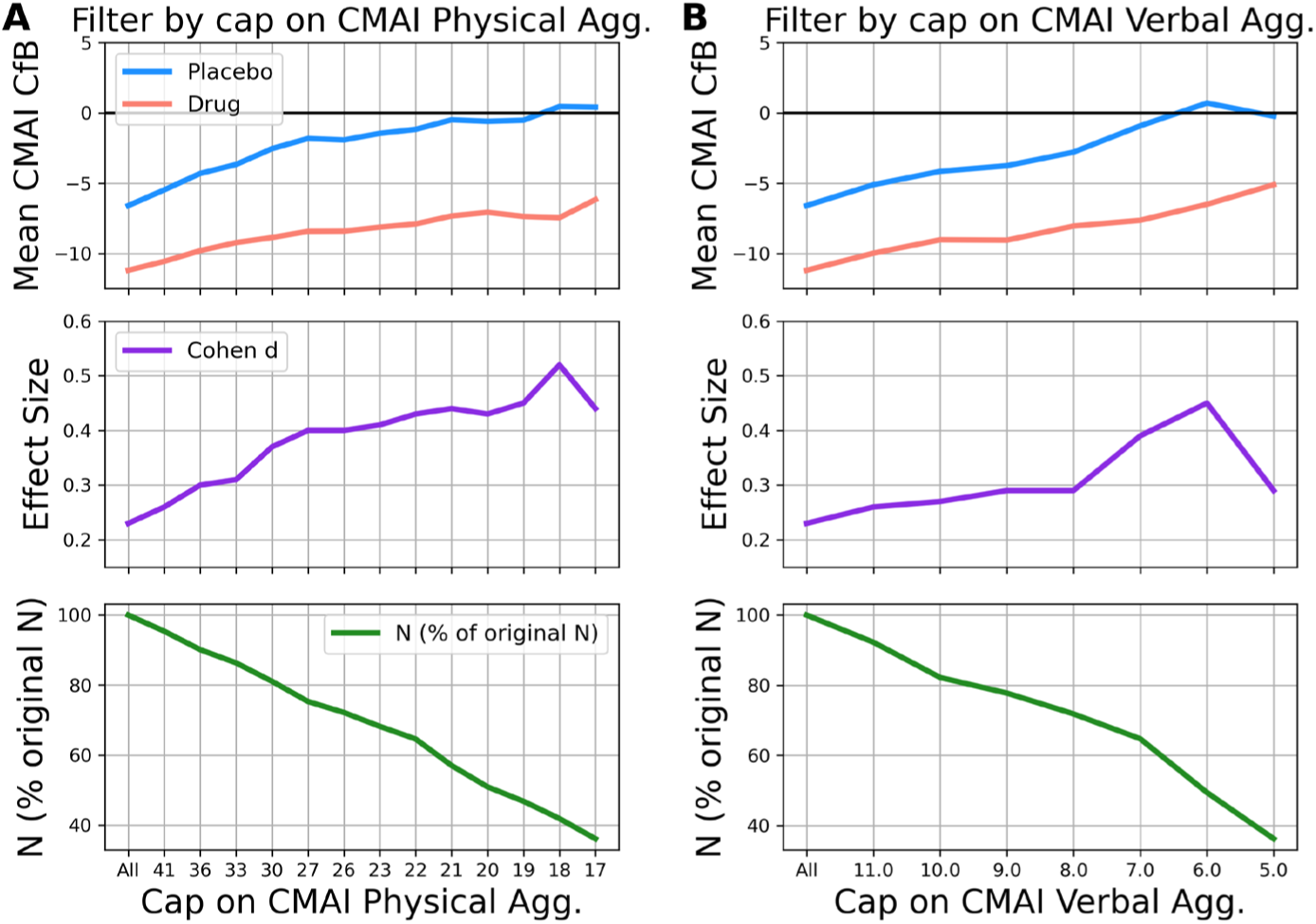
The effect within the combined training and validation datasets of baseline cap of CMAI physical aggressive (A) or verbal aggressive scores (B) on CMAI change scores in each arm (top), Cohen’s d effect size (middle) and percentage of the orginal participants who remained after the cap (bottom).

**Table S1.**
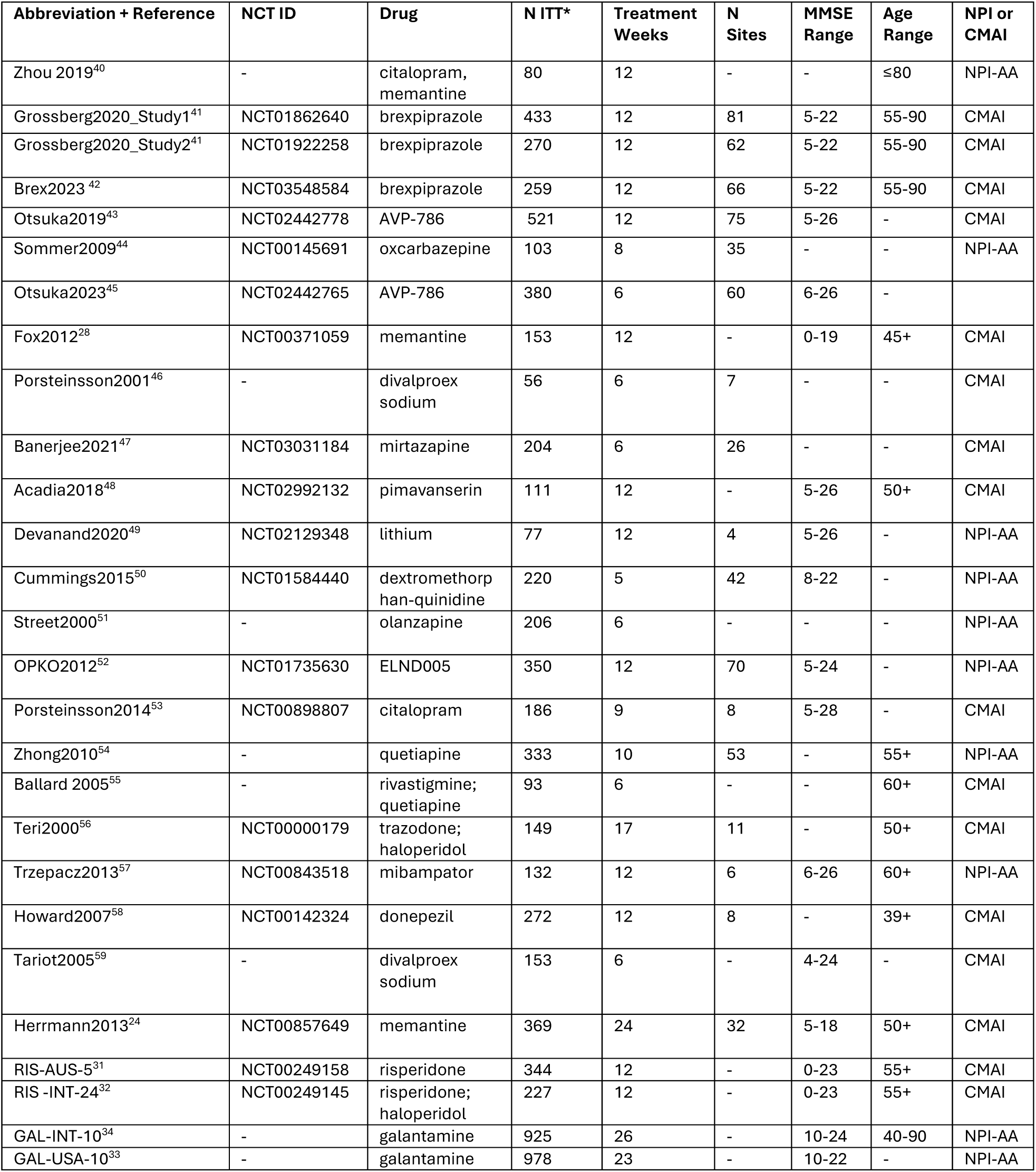
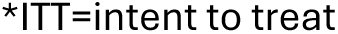

| Abbreviation + Reference | NCT ID | Drug | N ITT* | Treatment Weeks | N Sites | MMSE Range | Age Range | NPI or CMAI |
| --- | --- | --- | --- | --- | --- | --- | --- | --- |
| Zhou 2019<sup>40</sup> | - | citalopram, memantine | 80 | 12 | - | - | ≤80 | NPI-AA |
| Grossberg2020_Study1<sup>41</sup> | NCT01862640 | brexpiprazole | 433 | 12 | 81 | 5-22 | 55-90 | CMAI |
| Grossberg2020_Study2<sup>41</sup> | NCT01922258 | brexpiprazole | 270 | 12 | 62 | 5-22 | 55-90 | CMAI |
| Brex2023<sup>42</sup> | NCT03548584 | brexpiprazole | 259 | 12 | 66 | 5-22 | 55-90 | CMAI |
| Otsuka2019<sup>43</sup> | NCT02442778 | AVP-786 | 521 | 12 | 75 | 5-26 | - | CMAI |
| Sommer2009<sup>44</sup> | NCT00145691 | oxcarbazepine | 103 | 8 | 35 | - | - | NPI-AA |
| Otsuka2023<sup>45</sup> | NCT02442765 | AVP-786 | 380 | 6 | 60 | 6-26 | - |  |
| Fox2012<sup>28</sup> | NCT00371059 | memantine | 153 | 12 | - | 0-19 | 45+ | CMAI |
| Porsteinsson2001<sup>46</sup> | - | divalproex sodium | 56 | 6 | 7 | - | - | CMAI |
| Banerjee2021<sup>47</sup> | NCT03031184 | mirtazapine | 204 | 6 | 26 | - | - | CMAI |
| Acadia2018<sup>48</sup> | NCT02992132 | pimavanserin | 111 | 12 | - | 5-26 | 50+ | CMAI |
| Devanand2020<sup>49</sup> | NCT02129348 | lithium | 77 | 12 | 4 | 5-26 | - | NPI-AA |
| Cummings2015<sup>50</sup> | NCT01584440 | dextromethorphan-quinidine | 220 | 5 | 42 | 8-22 | - | NPI-AA |
| Street2000<sup>51</sup> | - | olanzapine | 206 | 6 | - | - | - | NPI-AA |
| OPKO2012<sup>52</sup> | NCT01735630 | ELND005 | 350 | 12 | 70 | 5-24 | - | NPI-AA |
| Porsteinsson2014<sup>53</sup> | NCT00898807 | citalopram | 186 | 9 | 8 | 5-28 | - | CMAI |
| Zhong2010<sup>54</sup> | - | quetiapine | 333 | 10 | 53 | - | 55+ | NPI-AA |
| Ballard 2005<sup>55</sup> | - | rivastigmine; quetiapine | 93 | 6 | - | - | 60+ | CMAI |
| Teri2000<sup>56</sup> | NCT00000179 | trazodone; haloperidol | 149 | 17 | 11 | - | 50+ | CMAI |
| Trzepacz2013<sup>57</sup> | NCT00843518 | mibampator | 132 | 12 | 6 | 6-26 | 60+ | NPI-AA |
| Howard2007<sup>58</sup> | NCT00142324 | donepezil | 272 | 12 | 8 | - | 39+ | CMAI |
| Tariot2005<sup>59</sup> | - | divalproex sodium | 153 | 6 | - | 4-24 | - | CMAI |
| Herrmann2013<sup>24</sup> | NCT00857649 | memantine | 369 | 24 | 32 | 5-18 | 50+ | CMAI |
| RIS-AUS-5<sup>31</sup> | NCT00249158 | risperidone | 344 | 12 | - | 0-23 | 55+ | CMAI |
| RIS -INT-24<sup>32</sup> | NCT00249145 | risperidone; haloperidol | 227 | 12 | - | 0-23 | 55+ | CMAI |
| GAL-INT-10<sup>34</sup> | - | galantamine | 925 | 26 | - | 10-24 | 40-90 | NPI-AA |
| GAL-USA-10<sup>33</sup> | - | galantamine | 978 | 23 | - | 10-22 | - | NPI-AA |

\*ITT=intent to treat

**Table S2.**

| Percentile in training data RIS-AUS-5 | CATE VALUE |
| --- | --- |
| 95 | 1.72 |
| 90 | 0.42 |
| 85 | -0.46 |
| 80 | -1.26 |
| 75 | -1.72 |
| 70 | -2.18 |
| 65 | -2.59 |
| 60 | -2.93 |
| 55 | -3.33 |
| 50 | -3.68 |
| 45 | -4.24 |
| 40 | -4.65 |
| 35 | -4.91 |
| 30 | -5.31 |
| 25 | -6.16 |
| 20 | -6.62 |
| 15 | -7.30 |
| 10 | -8.07 |
| 5 | -9.14 |

